# The Breadth of the Neutralizing Antibody Response to Original SARS-CoV-2 Infection is Linked to the Presence of Long COVID Symptoms

**DOI:** 10.1101/2023.03.30.23287923

**Authors:** Amanda M. Buck, Amelia N. Deitchman, Saki Takahashi, Scott Lu, Sarah A. Goldberg, Rebecca Hoh, Meghann C. Williams, Marian Kerbleski, Tyler-Marie Deveau, Sadie E. Munter, James Lombardo, Terri Wrin, Christos J. Petropoulos, Matthew S. Durstenfeld, Priscilla Y. Hsue, J. Daniel Kelly, Bryan Greenhouse, Jeffrey N. Martin, Steven G. Deeks, Michael J. Peluso, Timothy J. Henrich

## Abstract

**Background:** The associations between longitudinal dynamics and the breadth of SARS-CoV-2 neutralizing antibody response with various Long COVID (LC) phenotypes prior to vaccination are not known. The capacity of antibodies to cross neutralize a variety of viral variants may be associated with ongoing pathology and persistent symptoms.

**Methods:** We measured longitudinal neutralizing and cross-neutralizing antibody responses to pre- and post-SARS-CoV-2 Omicron variants in participants infected during the early waves of the COVID-19 pandemic, prior to wide-spread rollout of SARS-CoV-2 vaccines. Cross sectional regression models adjusted for various clinical covariates and longitudinal mixed effects models were used to determine the impact of the breadth and rate of decay of neutralizing responses on the development of Long COVID symptoms in general, as well as LC phenotypes.

**Results:** We identified several novel relationships between SARS-CoV-2 antibody neutralization and the presence of LC symptoms. Specifically, we show that, although neutralizing antibody responses to the original, infecting strain of SARS-CoV-2 were not associated with LC in cross-sectional analyses, cross-neutralization ID50 levels to the Omicron BA.5 variant approximately 4 months following acute infection was independently and significantly associated with greater odds of LC and with persistent gastrointestinal and neurological symptoms. Longitudinal modeling demonstrated significant associations in the overall levels and rates of decay of neutralization capacity with LC phenotypes. A higher proportion of participants had antibodies capable of neutralizing Omicron BA.5 compared with BA.1 or XBB.1.5 variants.

**Conclusions:** Our findings suggest that relationships between various immune responses and LC are likely complex but may involve the breadth of antibody neutralization responses.

**Summary:** SARS-CoV-2-specific antibody neutralization of Omicron BA.5 variant approximately 4 months following acute infection with wild-type virus prior to vaccination was independently and significantly associated with greater odds of distinct Long COVID phenotypes.

## INTRODUCTION

Many individuals experience post-acute sequelae of SARS-CoV-2 infection (PASC), which can affect quality of life and return to health [1-3]. The etiologic drivers of Long COVID (LC), a form of PASC defined by ongoing, often debilitating, symptoms, are poorly understood and likely involve multiple mechanisms [2, 4, 5]. Proposed mechanisms include aberrant autoreactive immune responses, microvascular dysregulation, and reactivation of latent human herpesviruses which may lead to the systemic inflammatory responses now identified in individuals with Long COVID compared to those who fully recovered [6-11]. Furthermore, there is growing evidence that SARS-CoV-2 subgenomic RNA and proteins are present in the tissues of at least a subset of immunocompetent individuals with LC [12-14]. Although those with persistent symptoms tend to have higher levels of SARS-CoV-2 Spike-specific antibody levels [10, 15-18], we and others have previously demonstrated that LC is associated with adaptive immune dysregulation and exhaustion [15, 18].

SARS-CoV-2 infection leads to rapid development of robust antibody responses, although neutralizing capacity wanes more quickly than total Spike IgG levels over time [17, 19-21]. A higher initial viral burden or persistence of viral antigens may lead to observed dysregulated immune phenotypes and higher antibody levels. However, there is a paucity of information regarding the associations between longitudinal dynamics or the breadth of the neutralizing antibody response with various LC phenotypes with some data showing that weaker antibody responses over time being associated with LC [22].

Recent pre-print data suggests that an expanded antibody response against the prior OC43 endemic coronavirus may be associated with Long COVID [23]. This suggests that the breadth of the response to initial infection may play an important role in the development of LC. Given that the rapid emergence of Omicron variants that evade neutralization result from infection from older SARS-CoV-2 strains (*e.g*., ancestral SARS-CoV-2, Alpha and Delta variants) as well as to COVID-19 vaccines [24, 25], there is strong interest in determining the relationship between the breadth and durability of the initial antibody responses and the presence of Long COVID symptoms. The rapid emergence of novel variants and increased incidence of reinfection also necessitates studies of longitudinal antibody responses following COVID-19 [26].

Here, we measured longitudinal neutralizing antibody responses to pre-Omicron strains and to subsequent Omicron variants in participants infected during the early waves of the COVID-19 pandemic, prior to their receiving SARS-CoV-2 vaccines. Cross sectional regression models adjusted for various clinical covariates and longitudinal mixed effects models were used to determine the impact of the breadth and rate of decay of neutralizing responses on the development of Long COVID symptoms in general, as well as distinct Long COVID symptom phenotypes.

## METHODS

### Clinical Cohort and Sample Collection

Participants were enrolled in the University of California, San Francisco (UCSF)-based Long-term Impact of Infection with Novel Coronavirus (LIINC) COVID-19 study (NCT04362150). The cohort design and procedures have been described in detail elsewhere [4]. Briefly, at each visit participants complete an interviewer-administered questionnaire querying the presence in the preceding 2 days of symptoms that are new since COVID-19 or worsened from pre-COVID baseline, prior to the collection of peripheral blood. This analysis included longitudinal measurements from 184 participants, including plasma samples collected between 1 and 4 months after initial symptom onset. All samples were collected prior to the participant having received any SARS-CoV-2 vaccination and a large majority were collected during the original SARS-CoV-2 wave (timing of sample collections here-maybe first and last date of collection), all prior to Omicron variant emergence. Phenotypic clusters were based on 32 participant-reported symptoms as previously described [4].

### PhenoSense SARS CoV-2 nAb Assay

The measurement of nAb activity using the PhenoSense SARS CoV-2 nAb Assay (Monogram Biosciences, South San Francisco, CA) was performed by generating HIV-1 pseudovirions that express the SARS CoV-2 Spike protein as previously described [20, 27-29]. The pseudovirus is prepared by co-transfecting HEK293 producer cells with an HIV-1 genomic vector that contains a firefly luciferase reporter gene together with a SARS CoV-2 Spike protein expression vector. Neutralizing antibody activity is measured by assessing the inhibition of luciferase activity in HEK293 target cells expressing the ACE2 receptor and TMPRSS2 protease following pre-incubation of the pseudovirions with serial dilutions of the serum specimen. ID50 values was generated for the original SARS-CoV-2 Spike protein as well as the following variants: Alpha (B.1.1.7), D614G mutant, Delta (B.1.617.2), Omicron BA.1 (B.1.1.529), Omicron BA.5 (B.1.1.529). A subset of samples stratified by negative or ID50s to the BA.5 strain were performed in an expanded pool consisting of pseudoviruses incorporating spike proteins from BA.2, BA.4.6, XBB.1.5 and BQ.1.1.

### Statistical analyses

Antibody data were generated blinded to participant information. Comparisons of ID50 values across comparator groups incorporated non-parametric Mann-Whitney or Freidman tests with Dunn correction for multiple comparisons using Prism v. 8 (GraphPad Software) and SPSS v. 29 (IBM). Adjusted P values reported in analyses involving multiple comparisons. For tabular data, two-tailed Fisher’s exact testing was performed on categorical data and two-tailed, non-parametric Mann-Whitney testing was performed on continuous variables (SPSS v. 29). Spearman Rank Correlation analysis was used to compare T cell, antibody and soluble markers of inflammation (Prism v. 8). For longitudinal analyses, linear mixed effects modeling was performed for neutralizing antibody ID50 (log transformed) in R (version 4.0.2) using lme4 package (version 1.1) with time and individual factors (*e.g*., age, sex, COVID-19 hospitalization, prior history of diabetes, prior autoimmune disease, body mass index >30, Long COVID symptoms) as predictors, and random effects based on participant.

Spearman’s correlation was performed in R to test for relationships between variant neutralizing antibody ID50 across all time points. Logistic regression models were performed on cross sectional data including the constant to identify independent associations between model factors and Long COVID outcomes using SPSS v. 29. Models included various demographic variables as well as either Log-transformed ID50 values or binary categorical indicators of ID50 values above a specific threshold as determined by sensitivity analyses.

### Study approval

All participants provided signed written informed consent prior to participation. The UCSF IRB approved the study.

## RESULTS

### Clinical cohort and participant demographics

To evaluate neutralizing responses in participants with prior COVID-19 and to assess relationships between these responses and the presence of Long COVID symptoms, we analyzed plasma samples collected from 184 participants with and without Long COVID symptoms across 384 timepoints for up to 4 months following acute infection. Participants had a median of 2 sample time points across all visits (ranging from 1 to 4). In general, participant visits occurred approximately 1 month, 2 months or 4 months following nucleic acid-confirmed SARS-CoV-2 infection. All specimens and symptom reports were timed from the day of initial COVD-19 symptom onset.

All participants were initially infected prior to emergence of the Delta strain (last date of infection was in March of 2021) and all but 7 samples across all time points were collected prior to the end of February 2021 (the time of vaccine availability to the general public in the Unites States). Long COVID was defined broadly as the presence of any symptom new or worsened since acute SARS-CoV-2 infection not clearly attributable to another cause. **Table 1** shows the clinical and demographic factors of participants grouped by the presence or absence of any Long COVID symptom. The group with Long COVID was enriched for women (58.4% vs 39.4%, P<0.05), those who had been hospitalized during acute COVID-19 (24.8% vs 19.7%, not significant), those with a history of pre-existing autoimmune disease (mainly thyroiditis, 10.6% vs 1.4%, P<0.05), persons self-reporting Latinx ethnicity (34.5% vs 19.7%, P<0.05), and those with higher body mass index (27.7 vs 26.0 kg/m^2^, P<0.05) (**Table 1**).

**Table 1.** Participant demographics, comorbidities, and clinical presentation in participants with and without post-acute sequelae of SARS-CoV-2 infection

|  | All Participants | No Long COVID | Long COVID<sup>a</sup> |
| --- | --- | --- | --- |
| N | 184 | 71 | 113 |
| Female Sex | 94 (51.1%) | 28 (39.4%) | 66 (58.4%)* |
| Age (median, IQR) | 44 (34, 53) | 43 (33, 53) | 45 (36, 54) |
| COVID-19 Hospitalization | 42 (22.8%) | 14 (19.7%) | 28 (24.8%) |
| Pre-Existing Medical Condition |  |  |  |
| Diabetes Mellitus | 20 (10.9%) | 11 (15.5%) | 9 (8.0%) |
| Autoimmune Disease | 13 (7.1%) | 1 (1.4%) | 12 (10.6%)* |
| Race/Ethnicity |  |  |  |
| Latinx | 53 (28.8%) | 14 (19.7%) | 39 (34.5%)* |
| White | 100 (54.3%) | 40 (56.3%) | 60 (53.1%) |
| Black/African American | 4 (2.2%) | 3 (4.2%) | 1 (0.9%) |
| Asian | 22 (12.0%) | 13 (18.3%) | 9 (8.0%) |
| American Indian/Native Alaskan | 1 (0.5%) | 0 (0%) | 1 (0.9%) |
| Pacific Islander/Native Hawaiian | 3 (1.6%) | 0 (0%) | 3 (4.2%) |
| Body Mass Index<sup>b</sup> | 26.7 (23.4, 31.2) | 26.0 (23.1, 28.8) | 27.7 (23.6, 32.9)* |

COVID-19 = coronavirus disease 2019; IQR = interquartile range
<sup>a</sup> Long COVID as defined as any persistent symptom new since COVID-19 onset at least 3 months following acute infection.
<sup>b</sup> n = 178 excluding missing values
\* P < 0.05 by two-sided Fisher's exact test for categorical data or by two-sided Mann-Whitney U test for continuous data.

### Breadth of antibody neutralization to the original infecting SARS-CoV-2 variant and subsequent viral variants

Using the PhenoSense assay (Monogram Biosciences), we measured the inhibitory serum dilutions at which 50% neutralization occurred (ID50) using pseudoviruses expressing SARS-CoV-2 Spike protein from the original strain (with which a majority of our participants were infected) and the following subsequent variants: Alpha (B.1.1.7), D614G mutant, Delta (B.1.617.2), Omicron BA.1 (B.1.1.529), and Omicron BA.5 (B.1.1.529). Overall, neutralization levels were highly variable between participants. Antibody neutralization responses were consistently highest to the original, infecting SARS-CoV-2 strain along with the Alpha, Beta, and Delta variants, with levels declining between 1 and 4 months following acute infection (**Figure 1A-C**). Although very low levels of cross-neutralization were observed with the Omicron BA.1 variant, a higher proportion of participants had antibodies able to neutralize Omicron BA.5 up to four months following initial presentation (the last study time point).

**Figure 1.**
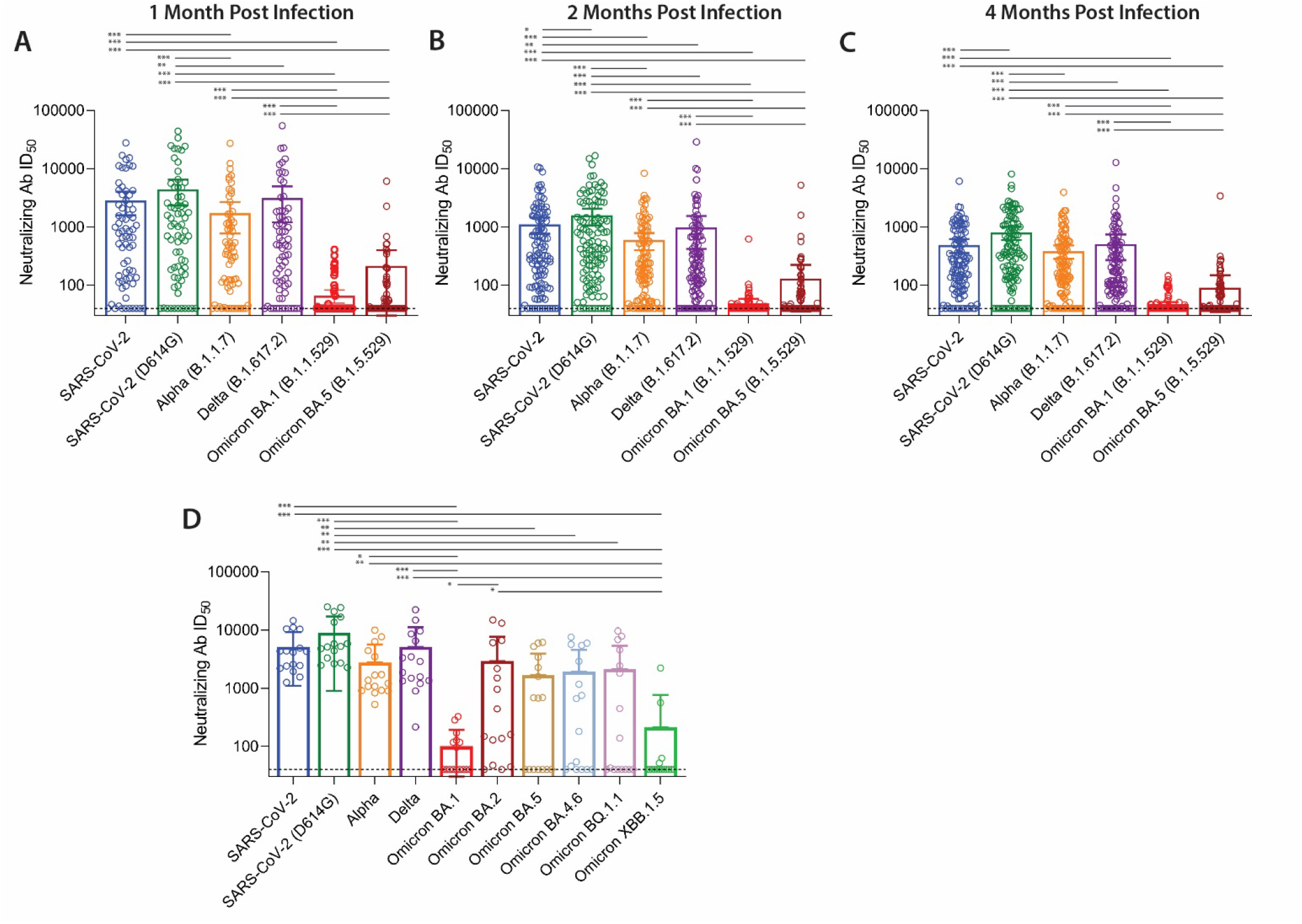
SARS-CoV-2 neutralization to the original infecting strain and cross-neutralization to subsequent viral variants. *Ex vivo* antibody neutralization of the original SARS-CoV-2 virus and subsequent variants approximately 1 month (**A**, N=69), 2 months (**B**, N=115), and 4 months (**C**, N=119) following acute infection for all participants. Subgroup analysis of cross-neutralization in an expanded Omicron sub variant panel (N=16) in a subset of participants that had samples across all timepoints with either high or negative neutralization to BA.5 (**D**). Most samples collected prior to SARS-CoV-2 vaccination and prior to the emergence of Delta and Omicron variants. Bars represent mean antibody (Ab) infectious dose 50% (ID50) values and 95% confidence intervals (* P<0.05, ** P<0.01, ***P<0.001 by two-sided Freidman test adjusted for multiple comparisons).

Across all time points, 12.5% of neutralizing titers were below the level of assay positivity to the original SARS-CoV-2 pseudovirus and 78.4% and 67.4% below the level of positivity for the BA.1 and BA.5 variant, respectively. Correlations of neutralization capacity to each SARS-CoV-2 variant are shown in **Supplementary Figure 1**.

We tested a subset of 16 participants including samples chosen with either high or low Omicron BA.5 responses across all timepoints on an expanded panel of Omicron sub variants to better understand the full breadth of cross-neutralization responses between ancestral and recent or current circulating strains as shown in **Figure 1D**. The expanded neutralization panel included BA.4.6, BQ.1.1 and XBB.1.5. Similar neutralization titers were observed between BA.5, BA.4.6, and BQ1.1, whereas XBB.1.5 responses most closely resembled BA.1 responses, which were overall low or negative across participants. BA.2 responses were more evenly distributed across a range of neutralization ID50s compared to the other Omicron sub variants.

### Decay of variant-specific SARS-CoV-2 antibody neutralization by clinical phenotype

Leveraging mixed effects modeling approaches, we analyzed neutralizing responses over time by clinical and demographic characteristics, including age, sex, hospitalization during acute infection, body mass index (BMI), and a pre-existing history of diabetes mellitus for the most clinically relevant variants in our study population (ancestral SARS-CoV-2, Delta and Omicron sub variants; **Figure 2**). Overall, antibody neutralization ID50 decreased over time for all strains (p<0.01) with the exception of the Omicron BA.1 variant, (p=0.16) for which initial levels were substantially lower than to the other variants. When stratifying by age greater than 50 years, we identified no difference in antibody neutralization to all variants tested across all time points in the mixed effects models. In contrast, male sex was associated with higher viral neutralization for original SARS-CoV-2, Delta, and Omicron BA.5 variants, but not for Omicron BA.1, across all time points. These observed sex-based differences were similar between original and Delta variants, each approximately 0.41 and 0.33 log_10_ higher, respectively, observed in males compared to females recovering from COVID-19.

**Figure 2.**
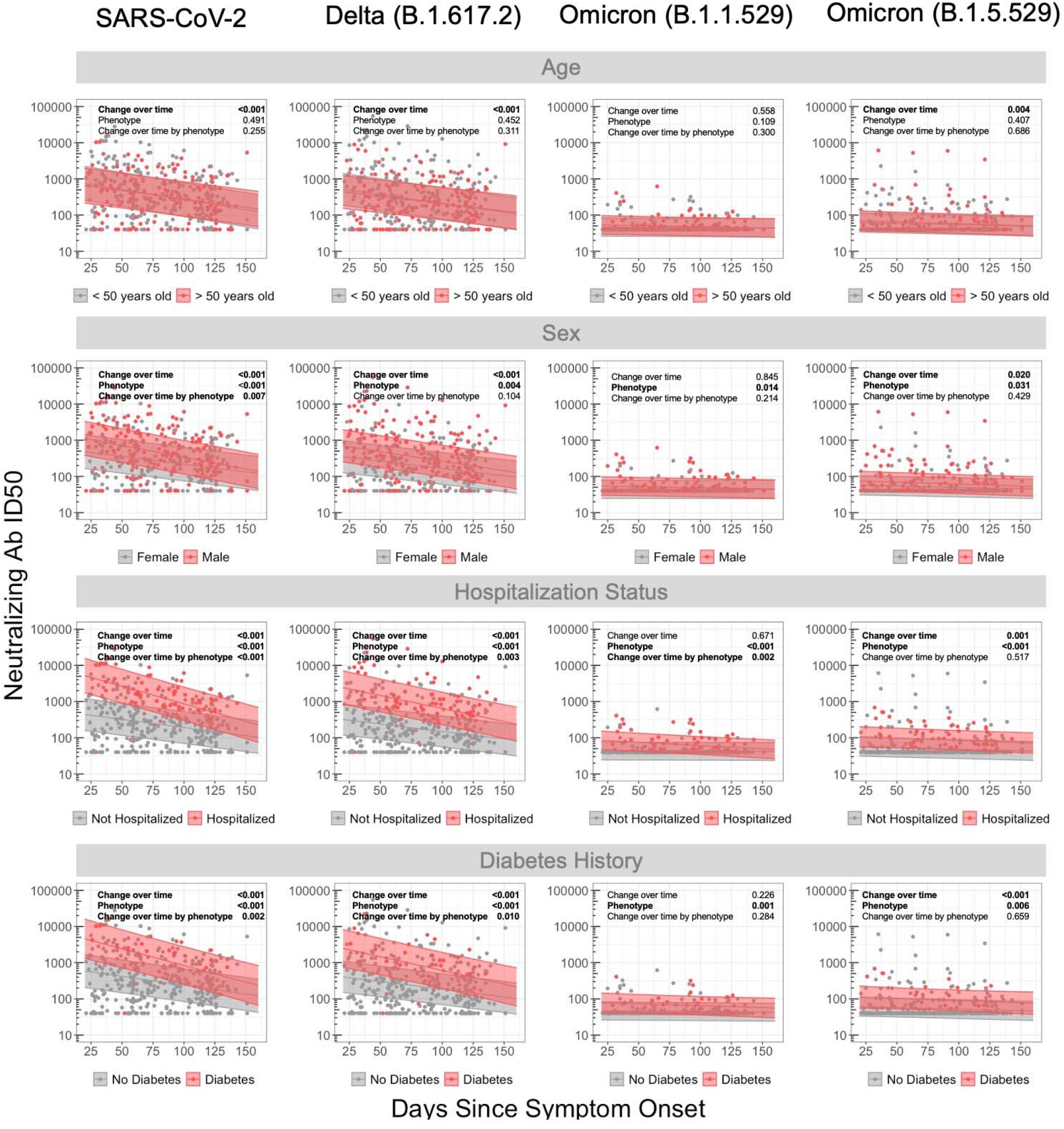
Longitudinal analysis of antibody neutralization by demographics, clinical factors and SARS-CoV-2 variant. Mixed-linear regression model with four covariates (*i.e*. age, sex, hospitalization status, and prior diabetes history) for different variants: SARS-CoV-2, B.1.617.2, B.1.1.529 and BA.4/5. P-values denote if a significant difference was observed for change in antibody neutralization over time (Change over time), between subgroup (Phenotype, e.g., no diabetes versus diabetes), and difference in change over time by subgroup (Change over time by phenotype, *e.g*. difference in slope of decline in antibody levels dependent on diabetes versus no diabetes). Shaded region represents 95% confidence intervals around the median line.

For those hospitalized during acute infection, neutralization levels were significantly higher across all strains with the exception of BA.1. The magnitude of the difference in responses for those hospitalized versus not hospitalized was diminished with those strains with lower overall responses: 1.18 and 0.94 log_10_ higher responses for the original virus and Delta variant versus 0.33 and 0.28 log_10_ higher for omicron BA.1 and BA.5 responses, respectively. Individuals with pre-existing diabetes also had differential neutralization across variants, with higher initial levels and longitudinal levels over all time points, and showing a more rapid decline over time compared to those without diabetes. The magnitude of wild-type SARS-CoV-2 responses for those with a BMI >30 was higher across all time points for the original and delta viral strains (All P<0.001) and declined more rapidly for the original Delta and Omicron BA.5 variants (All P>.012; **Supplemental Figure 2**). In contrast, neutralization ID50s from participants with a pre-existing history of diabetes mellitus were overall lower across all time points for the original SARS-CoV-2 (All P <0.05) and ID50s to the original strain and Delta variant declined less rapidly (P<0.05).

### Breadth of the neutralizing antibody responses is associated with increased odds of Long COVID

In order to determine whether neutralization capacity was related to Long COVID, we performed logistic regression modeling with either the presence of any Long COVID symptom at a given sample time point or with specific Long COVID symptom phenotype (neurocognitive, cardiopulmonary, gastrointestinal, musculoskeletal and fatigue) at the three main collection time points: 1 month (N=69; median 33 days), 2 months (N=115; median 59 days) and 4 months (N=119; median 120 days) following acute infection. Data from only one time point per participant within each time period was included to avoid oversampling of specific individuals. Specifically, the sample time closest to 30 days within a 21-45 day window, 60 days within a 56-75 day window, and 120 days within a 100-150 day window were included. Factors included in the first model (**Figure 3A**) included the neutralizing antibody ID50 (continuous variable), prior hospitalization during acute COVID-19, female sex, and age greater than 50 years of age. Overall, there were no significant differences between neutralization ID50 to any strain and the presence of Long COVID in general or any specific Long COVID phenotype at 1 and 2 months following acute infection (all P>0.05). As shown in **Figure 3A**, the neutralization ID50 of the ancestral SARS-CoV-2 (the infecting strain in this study population), as well as Alpha and Delta variants, were not significantly associated with the presence of any Long COVID symptom or specific Long COVID phenotype approximately 4 months after acute infection. However, cross-neutralization ID50s to Omicron BA.5 were significantly and positively associated with neurocognitive and gastrointestinal symptoms (*i.e*. higher odds of having symptoms within these phenotypes). There were no significant associations between BA.5 neutralization ID50 and fatigue and cardiopulmonary symptoms or as shown in **Supplementary Figure 3**). Regression analyses including ID50s to both wild-type SARS-CoV-2 and Omicron BA.3 were also performed as in Supplementary **Figure 4A**. Including ID50s to both wild type and BA.5 strains led to similar results, with cross-neutralization to Omicron BA.5 being significantly associated with having any Long COVID symptom and neurocognitive symptoms, whereas ancestral SARS-CoV-2 ID50 were not significantly associated with Long COVID or any symptoms cluster.

**Figure 3.**
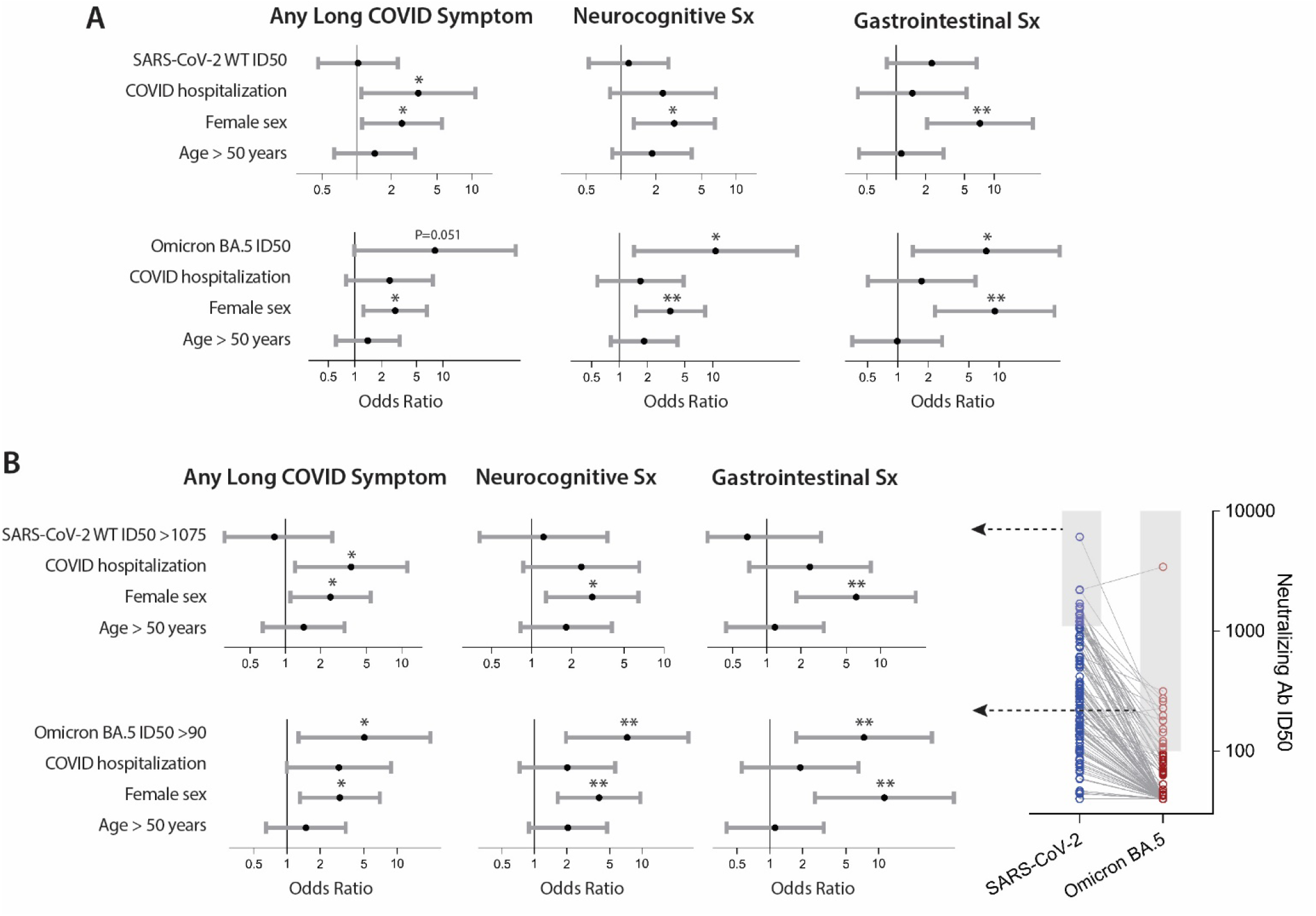
Association between SARS-CoV-2 neutralization, hospitalization during acute infection and demographic factors and the odds of experiencing Long COVID approximately four months following acute COVID-19. The top panel shows odds ratios (points) and 95% confidence intervals (bars) for each variable included in logistic regression models using continuous neutralization ID50 values for assays using the original and Omicron BA.5 pseudoviruses (**A**). The bottom panel shows odds ratios and 95% confidence intervals of developing PASC or specific PASC phenotypes for logistic regression models incorporating a binary variable indicating if a sample had a neutralization ID50 in the top 15% of the cohort to either original SARS-CoV02 or Omicron BA.5 pseudovirus (**B**). * P<0.05, ** P<0.01 from covariate adjusted logistic regression.

Female sex was positively and significantly associated with an increased odds of Long COVID in models including both the original infecting virus and cross-neutralization to the Omicron BA.5 variant. Hospitalization for acute COVID-19 was only significantly associated with Long COVID in the model incorporating the original SARS-CoV-2 strain pseudovirus. A pre-existing history of diabetes mellitus or autoimmune disease were included in subsequent regression models but were not significantly associated with Long COVID outcomes and did not influence significance of other factors.

To further test the relationship between cross-neutralization of BA.5 pseudoviruses with the development of Long COVID symptoms, we performed binary logistic regression including only the top 15% of neutralization ID50s in both the original SARS-CoV-2 and BA.5 variant based on results from a sensitivity analysis to evaluate the influence of the samples with robust cross-neutralization of BA.5 as shown in **Figure 3B**. Consistent with the above analyses, there were no significant associations between the top neutralization responders to the original virus and PASC or PASC phenotype, whereas presence of robust cross-neutralization to Omicron BA.5 was significantly associated with a higher odds of any PASC symptom and neurocognitive and gastrointestinal PASC phenotypes. We repeated regression analysis with ancestral wild-type and Omicraon BA.5 in the same model and results were similar with high cross-neutralization to BA.5 being significantly associated with any Long COVID symptom and neurocognitive and gastrointestinal symptoms clusters (**Supplementary Figure 4B**).

### Decay of SARS-CoV-2-specific antibody neutralization over time by Long COVID phenotype

Finally, we assessed neutralizing antibody responses against the original infecting SARS-CoV-2 strain by individual Long COVID phenotype (*i.e*. non mutually exclusive symptom cluster) compared to those without any Long COVID symptoms or those with or without symptoms but not within the specific Long COVID symptom cluster (**Figure 4A & B**, respectively). Overall, differences in levels across all time points or changes over time were similar, with high interpatient variation in neutralization ID50s observed. Nonetheless, we found that those with gastrointestinal and cardiopulmonary symptoms had 0.27 and 0.43 log_10_ higher neutralization ID50 compared to those without any Long COVID symptom across all data points over time (P=0.04 and <0.001, respectively; **Figure 4A**). Decay in neutralization ID50 was faster (*i.e*. more negative slope in mixed linear effects model) in participants with cardiopulmonary and musculoskeletal symptoms compared to those without any symptoms (P=0.01 and 0.047, respectively). Compared to those with or without persistent symptoms, but no symptoms in the specified phenotype cluster, those with cardiopulmonary or musculoskeletal symptoms were overall higher across all time points (both P<0.001), and those with musculoskeletal, cardiopulmonary and neurocognitive symptoms declined more rapidly than those without those specific symptoms (all P < 0.05; **Figure 4B**).

**Figure 4.**
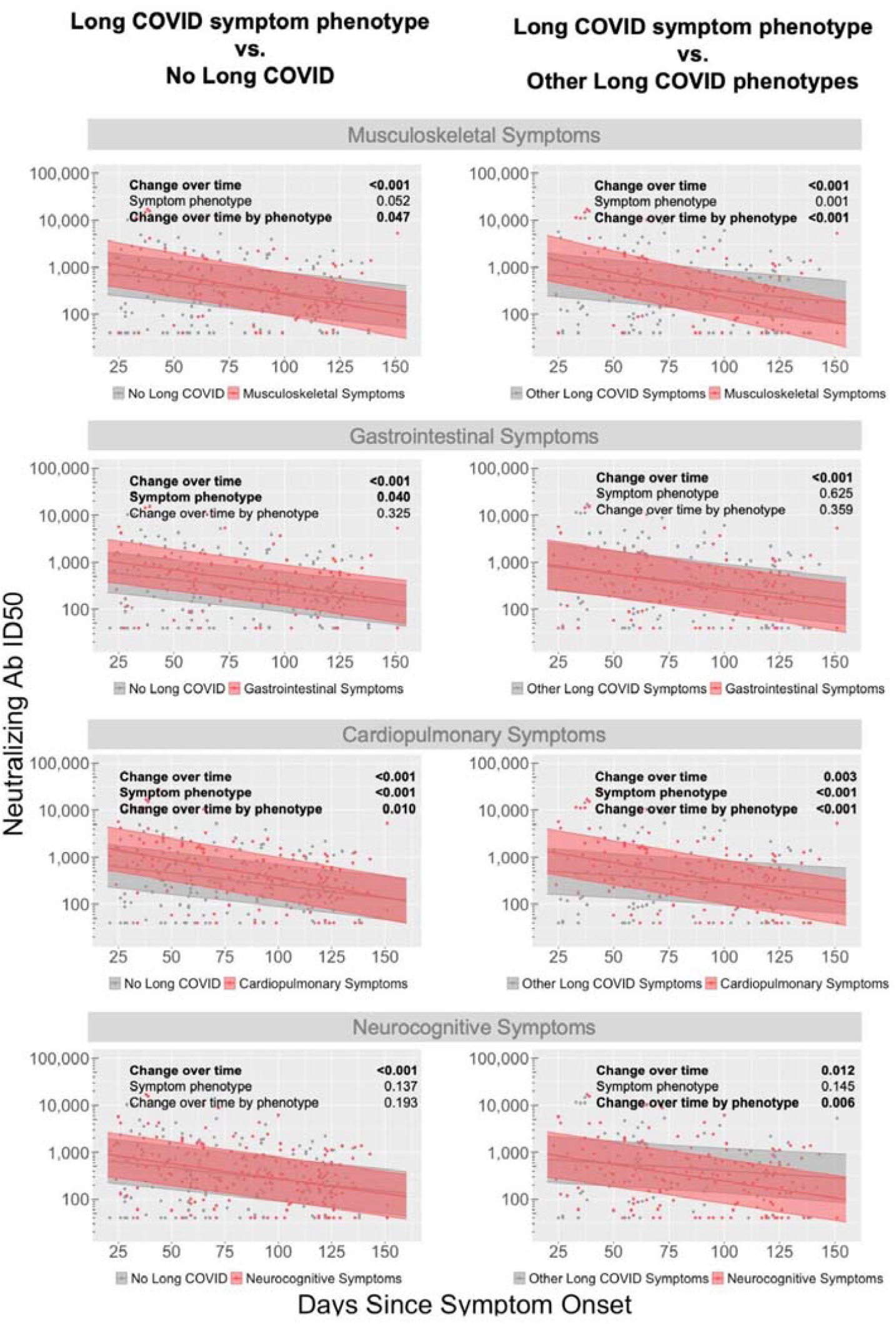
Differential decay of SARS-CoV-2-specific neutralizing antibody responses among Long COVID Symptom Phenotypes. Longitudinal decay of antibody responses compared between participants with Long COVID Symptom Phenotype (e.g. musculoskeletal, gastrointestinal, cardiopulmonary, neurocognitive) **a)** versus those without Long COVID (left panels) or **b)** versus those with other Long COVID Symptom Phenotypes (right panels). Each panel includes p-values for: the decay across all participants (“change over time”), differences in antibody neutralization for those with Long COVID phenotype across all timepoints (Symptom Phenotype), and differences in change over time with a given Long COVID phenotype (Change over time by phenotype). Shaded region represents 95% confidence intervals around the median line.

## DISCUSSION

In this longitudinal study of a well-characterized cohort of people recovering from COVID-19 during the early waves of the pandemic prior to emergence of Delta and subsequent variants and availability of vaccines, we identified several novel relationships between SARS-CoV-2 antibody neutralization and the presence of Long COVID symptoms. First, we show that, although neutralizing antibody responses to the original, infecting strain of SARS-CoV-2 were not associated with Long COVID in cross-sectional analyses, cross-neutralization ID50 levels to the Omicron BA.5 variant approximately 4 months following acute infection were independently and significantly associated with greater odds of Long COVID in general and specifically with persistent gastrointestinal and neurological symptoms. These data suggest that a broad antibody response to subsequent viral variants may predict emergence of various LC symptoms. Supporting this finding, a recent preprint suggested that people with LC may have an expanded antibody response against the prior OC43 endemic Coronavirus [23]. The researchers also demonstrated more avid IgM responses and inflammatory OC43 S2-specific Fc-receptor binding responses but weaker Fc-receptor binding to SARS-CoV-2 [23]. Whether or not the association between breadth of response and LC is due to or related to processes such as autoreactive antibody formation warrants further investigation.

Interestingly, we observed that a higher proportion of people infected early during the pandemic had antibodies capable of neutralizing Omicron BA.5 compared with BA.1, to which very few participants demonstrated any cross-neutralization response. The reason for this is not known, but several amino acid mutations in the receptor binding domain (RBD) of SARS-CoV-2 Spike protein in the BA.1 strain reverted to wild-type in the BA.5, such as G446S, Q493R and G496R [19, 30]. The BA.5 variant also had reversion of amino acid mutations or insertions in non RBD areas of Spike to pre BA.1 variants (*e.g*. L981F, ins214EPE, T95I) [19, 30]. In the sub-analysis of samples from 16 participants stratified by the highest and lowest BA.5 neutralization titers, we also observed consistently higher cross-neutralization with BA.2, BA.4.6 and BQ.1.1 sub variants. In contrast, lower neutralization ID50s to XBB.1.5 pseudovirus were observed, similar to BA.1 neutralization. Whether or not there are any clinical implications of increased cross-neutralization in those infected early in the pandemic with subsequent Omicron variants is not known. It is also not clear if infection with Omicron strains leads to increased or decreased risks of Long COVID, but it will be very difficult to identify variant-specific effects in the current era of widespread and variable vaccination, reinfection, and antiviral treatment uptake.

We also evaluated the longitudinal relationships between antibody neutralization responses, various clinical factors, and Long COVID phenotypes. The mixed effects models allowed us to determine differences between these variables across all data points over time (including multiple time points for each individual) and changes (*i.e*. decay) over time. These analyses revealed relationships between neutralization responses that were not observed in the cross-sectional analyses, such as significantly higher SARS-CoV-2 neutralizing responses to the original infection SARS-CoV-2 across all time points for those with gastrointestinal and cardiopulmonary Long COVID symptoms. Regardless of the overall higher levels, a faster decay of neutralizing ID50 was observed for these phenotypic clusters, in addition to those with musculoskeletal symptoms. Of note, neutralization titers decay more rapidly on a whole than common epitope antibody responses which have been associated with various PASC symptoms in longitudinal analyses [16, 17, 20], highlighting importance of temporal immune dynamics in the study of this condition. Together, these and the cross-sectional data suggest that an overall higher neutralization response that wanes more quickly, or ones that remain broad over time, are associated with LC. While the causes of these dynamics are unknown, one possibility is that persistent SARS-CoV-2 antigen presentation in tissues, which has been proposed as a potential mechanism of Long COVID, may lead to overall higher antibody neutralization over time, and potentially to a broader response to subsequent variants. While speculative, our findings suggest that relationships between various immune responses and Long COVID are likely complex, and different approaches to data analyses may yield different, but complementary information.

Strengths of this study include the use of highly characterized samples from the pre-vaccine and pre-Omicron era, before reinfections became common. This allowed for a more straightforward analysis of neutralization dynamics in the absence of these complex confounding factors. In addition, both those with and without Long COVID were recruited and assessed in an identical manner, addressing potential biases that might occur when comparison groups are derived from different cohorts as has been common in studies of Long COVID. As in similar analyses where we have been able to evaluate mechanisms according to distinct Long COVID phenotypes [9], we leveraged our high degree of symptom characterization to analyze different case definitions of Long COVID. This approach is informative, especially since the case definition remains controversial and it is possible that different phenotypes are driven by different mechanisms. Limitations of the study include a lack of participants infected with more recent variants, preventing us from extending our observations into more recent waves of the pandemic. The cohort was a convenience sample, and although this allows for valid inferences regarding Long COVID biology comparing people with and without the phenotype of interest within the cohort, extrapolation to all individuals with prior COVID-19 must be done cautiously. The neutralization assay used Spike protein pseudoviruses rather than intact, replication-competent virus, but these pseudovirus assays have been shown to have comparable results in several studies [27, 29, 31-33]. Nonetheless, we believe that these results suggest at least one potential contributor to Long COVID, although more work will be necessary to validate these observations in other cohorts, including those derived from later waves of the pandemic in the setting of vaccination or reinfection.

## Data Availability

All data produced in the present study are available upon reasonable request to the authors.

## Acknowledgements

We are grateful to the study participants and their medical providers. We acknowledge current and former LIINC clinical study team members Tamara Abualhsan, Andrea Alvarez, Khamal Anglin, Urania Argueta, Mireya Arreguin, Kofi Asare, Melissa Buitrago, Monika Deswal, Nicole DelCastillo, Emily Fehrman, Halle Grebe, Heather Hartig, Yanel Hernandez, Beatrice Huang, Raushun Kirtikar, Monica Lopez, Michael Luna, Lynn Ngo, Enrique Martinez Ortiz, Antonio Rodriguez, Justin Romero, Dylan Ryder, Ruth Diaz Sanchez, Matthew So, Celina Chang Song, Viva Tai, Alex Tang, Cassandra Thanh, Fatima Ticas, Leonel Torres, Brandon Tran, Daisy Valdivieso,and Deepshika Varma; and LIINC laboratory team members Joanna Donatelli, Jill Hakim, Nikita Iyer, Owen Janson, Brian LaFranchi, Christopher Nixon, Isaac Thomas, and Keirstinne Turcios. We thank Jessica Chen, Aidan Donovan, Carrie Forman, and Rania Ibrahim for assistance with data entry and review. We thank the UCSF AIDS Specimen Bank for processing specimens and maintaining the LIINC biospecimen repository. We are grateful to Elnaz Eilkhani and Monika Deswal for regulatory support. We are also grateful for the contributions of LIINC leadership team members: Isabelle Rodriguez-Barraquer, and Rachel Rutishauser.

## SUPPLEMENTARY MATERIALS

**Supplementary Figure 1.**
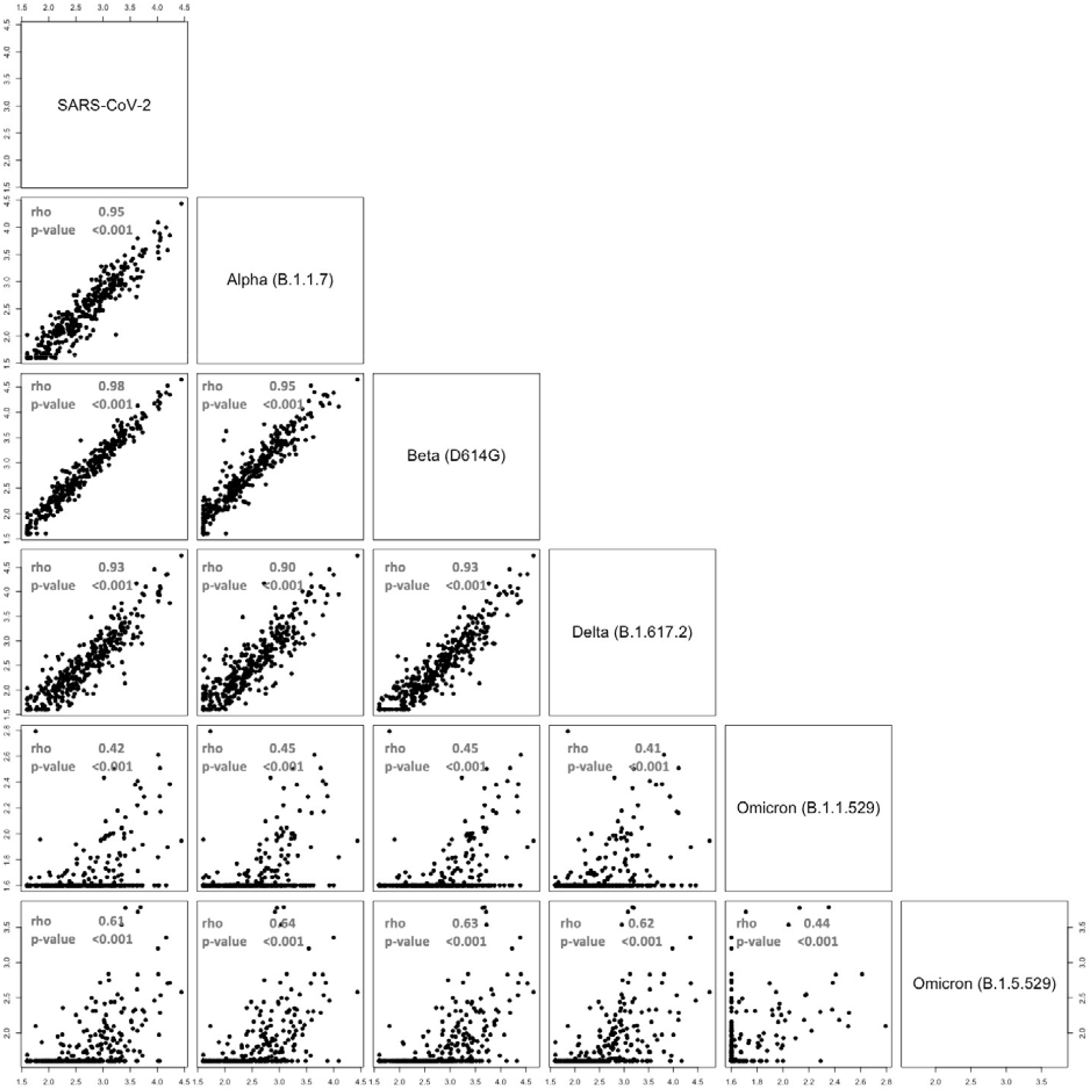
Correlation Matrix of Neutralization ID50s For SARS-CoV-2 variants tested. Data across all time points are shown in the matrices. R and P values from non-parametric Spearman rank correlation analyses.

**Supplementary Figure 2.**
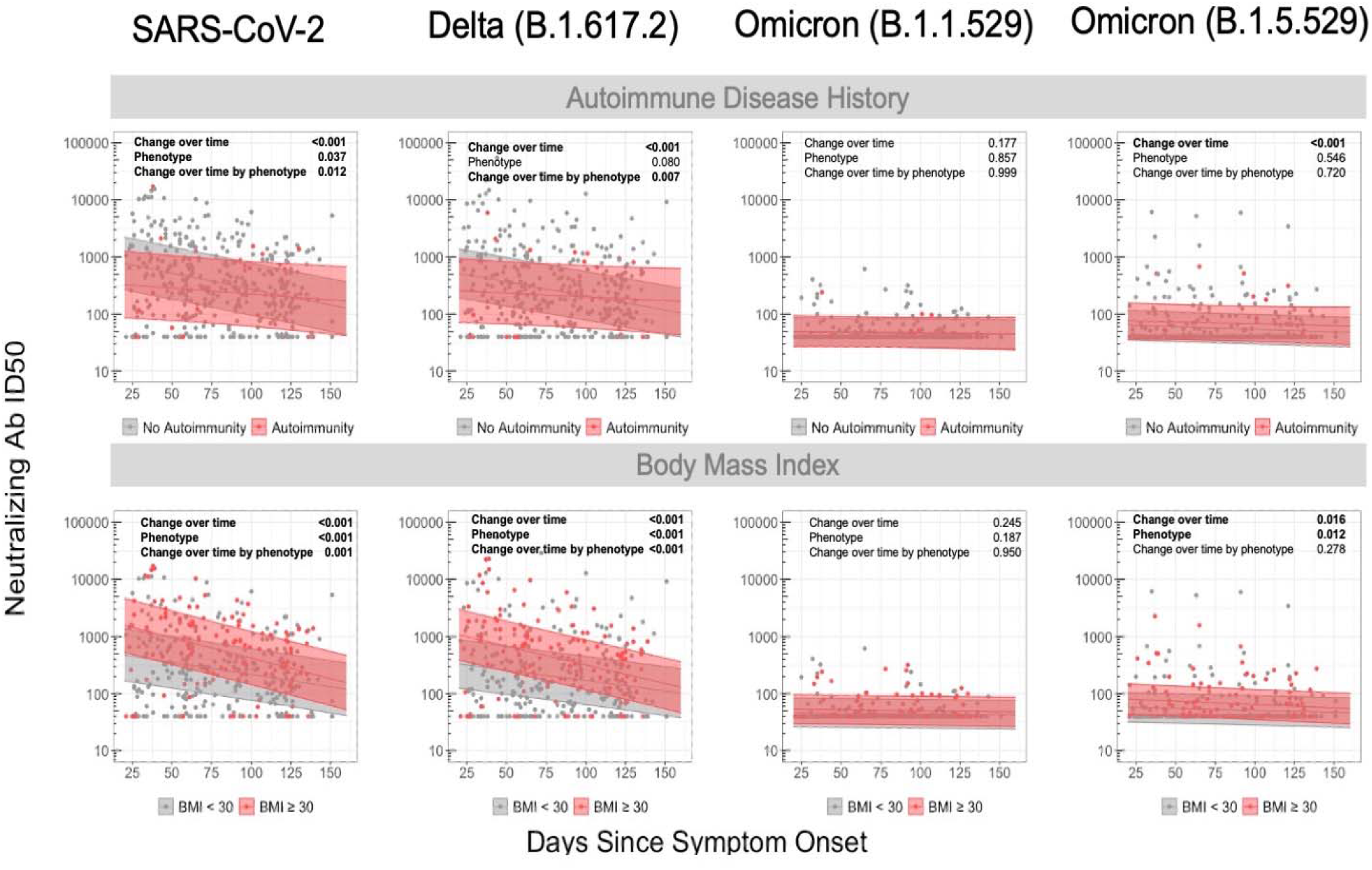
Longitudinal analysis of antibody neutralization by clinical factors and SARS-CoV-2 variant. Mixed-linear regression model with two covariates (body mass index ad autoimmune disease history) for different variants: SARS-CoV-2, B.1.617.2, B.1.1.529 and BA.4/5. P-values denote if a significant difference was observed for change in antibody neutralization over time (Change over time), between subgroup (Phenotype, *e.g*., BMI ≥ 30), and difference in change over time by subgroup (Change over time by phenotype). Shaded region represents 95% confidence intervals around the median line.

**Supplementary Figure 3.**
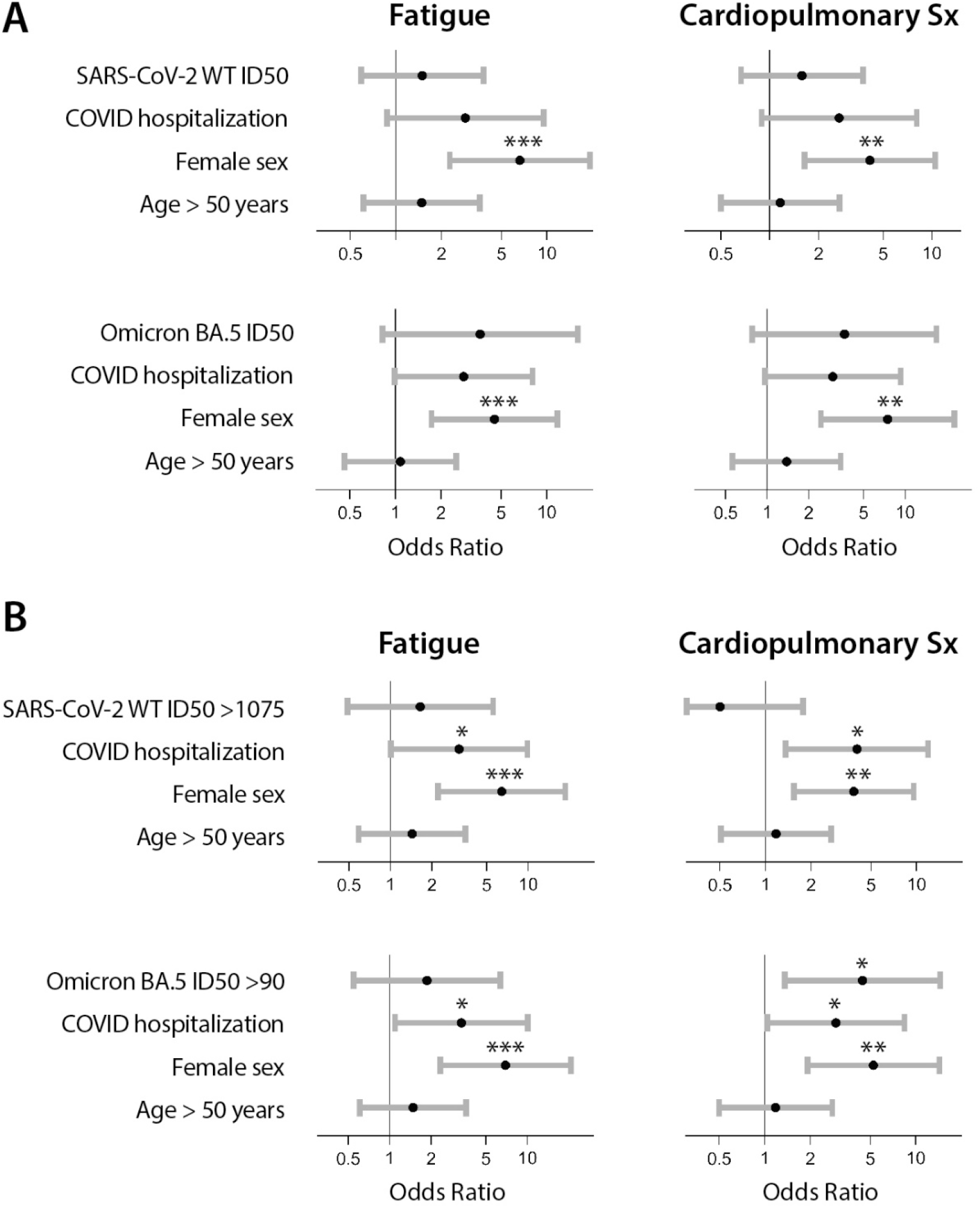
Association between SARS-CoV-2 neutralization, hospitalization during acute infection and demographic factors and the odds of experiencing fatigue or cardiopulmonary symptoms approximately four months following acute COVID-19. The top panel shows odds ratios (points) and 95% confidence intervals (bars) for each variable included in logistic regression models using continuous neutralization ID50 values for assays using the original and Omicron BA.5 pseudoviruses (**A**). The bottom panel shows odds ratios and 95% confidence intervals of developing PASC or specific PASC phenotypes for logistic regression models incorporating a binary variable indicating if a sample had a neutralization ID50 in the top 15% of the cohort to either original SARS-CoV02 or Omicron BA.5 pseudoviruses (**B**). *P<0.05, **P<0.01, ***P<0.001 from covariate adjusted logistic regression.

**Supplementary Figure 4.**
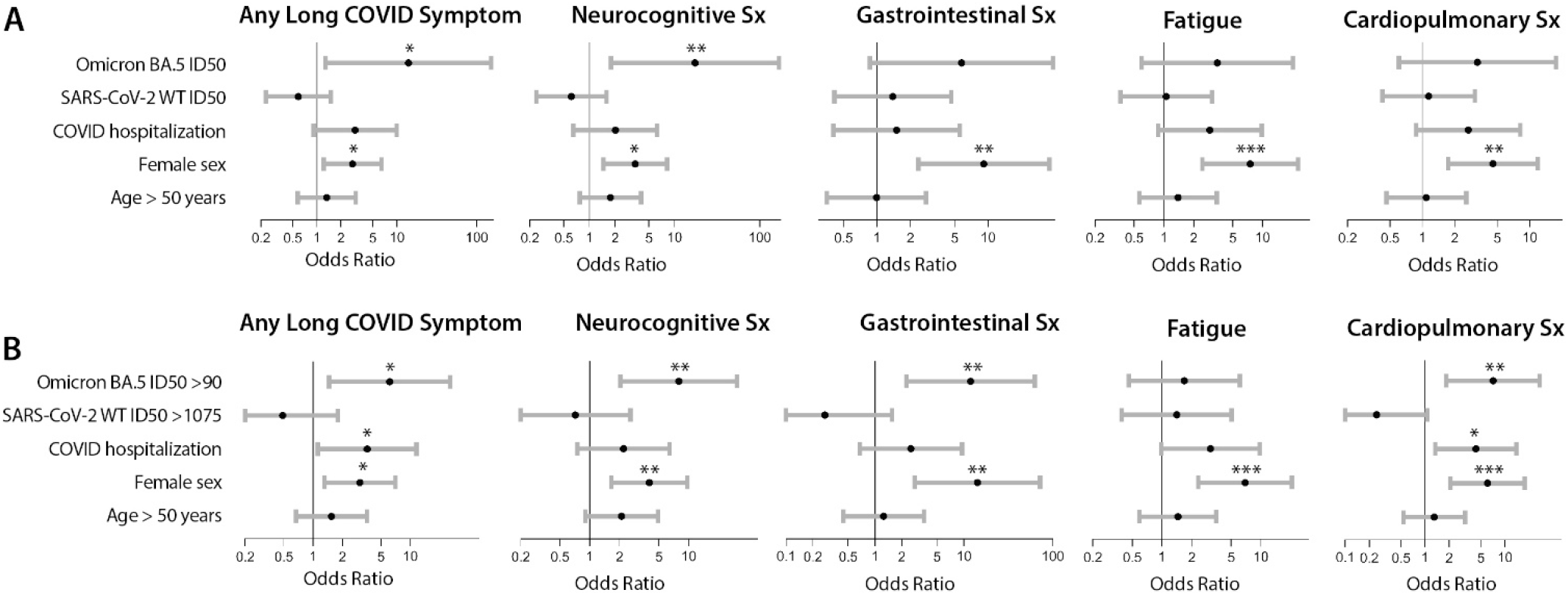
Association between SARS-CoV-2 neutralization, hospitalization during acute infection and demographic factors and the odds of experiencing fatigue or cardiopulmonary symptoms approximately four months following acute COVID-19. The top panel shows odds ratios (points) and 95% confidence intervals (bars) for each variable included in logistic regression using continuous neutralization ID50 values for assays using the original and Omicron BA.5 pseudoviruses in the same model (**A**). The bottom panel shows odds ratios and 95% confidence intervals of developing PASC or specific PASC phenotypes for logistic regression incorporating a binary variable indicating if a sample had a neutralization ID50 in the top 15% of the cohort to the original SARS-CoV02 and Omicron BA.5 pseudoviruses (**B**). *P<0.05, **P<0.01, ***P<0.001 from covariate adjusted logistic regression.

